# Enhancing gout management by creating a register using automated queries in electronic health records

**DOI:** 10.1101/2024.03.08.24303964

**Authors:** Nils Bürgisser, Denis Mongin, Samia Mehouachi, Clement P. Buclin, Romain Guemara, Pauline Darbellay Farhoumand, Olivia Braillard, Kim Lauper, Delphine S. Courvoisier

**Affiliations:** Division of Rheumatology, Department of Medicine, Geneva University Hospitals, Geneva, Switzerland; Division of General Internal Medicine, Department of Medicine, Geneva University Hospitals, Geneva, Switzerland; Division of Primary Care Medicine, Geneva University Hospitals, Geneva, Switzerland; Quality of Care Unit, Geneva University Hospitals, Geneva, Switzerland; Faculty of medicine, University of Geneva, Geneva, Switzerland; Geneva Center for Inflammation research, Department of Medicine, Faculty of Medicine, University of Geneva, Geneva, Switzerland

## Abstract

**Objective:** To develop an automatic gout register to improve gout management.

**Methods:** We analysed the electronic health records (EHR) of all patients >18 years old from a tertiary academic hospital (2013-2022) based on six criteria: International Classification of Diseases 10 (ICD-10) gout diagnosis, urate-lowering therapy (ULT) prescription, uric acid crystal in joint aspiration and gout-related terms in problem lists, clinical or imaging reports. We assessed the positive and negative predictive value (PPV and NPV) of the query by chart reviews.

**Results:** Of 2,110,902 out- and inpatients, 10,289 had at least one criterion for gout. The combination of joint aspiration OR diagnostic in the problem list OR ≥ 2 other criteria created a register of 5,138 patients, with a PPV of 92.4% (95%CI: 88.5 to 95.0), and an NPV of 94.3% (95%CI: 91.9 to 96.0). PPV and NPV were similar amongst outpatients and inpatients. Incidence was 2.9 per 1000 person-year and dropped by 30% from the COVID-19 pandemic onward. Patients with gout were on average 71.2 years old (SD 14.9), mainly male (76.5%), overweight (69.5%) and polymorbid (mean number of comorbidities of 3, IQR 1-5). More than half (57.4%) had received a urate lowering treatment, 6.7% had a gout that led to a hospitalisation or ≥2 flares within a year, and 32.9% received a rheumatology consultation.

**Conclusion:** An automatic EHR-based gout register is feasible, valid and could be used to evaluate and improve gout management. Interestingly, the register uncovered a marked underdiagnosis or underreporting of gout since the COVID-19 pandemic.

*Key messages:* What is already known on this topic? - Gout is the most prevalent inflammatory arthritis, but it remains undertreated despite affordable and effective treatment options.
- Quantifying this undertreatment and detecting its causes and risk factors to pilot quality improvement initiative requires an extensive register of gout patients. What this study adds? - This is the first automatic EHR-based gout register, allowing frequent, inexpensive, and sustainable updates.
- The automated queries show high positive and negative predictive values to identify gout patients. How this study might affect research, practice or policy? - This register can facilitate the assessment of the adequacy of gout management and the monitoring of quality indicators following improvement projects, or change in policies
- It provides an easy platform for cohort studies or adaptive trials
- Its methodology is reproducible, facilitating the establishment of gout or other disease registers within different EHR systems

## Introduction

Gout is a chronic accumulation of uric acid crystal in joints and surrounding tissues. It manifests as a disease continuum, ranging from asymptomatic monosodium urate crystal deposition to acute debilitating joint flares, which lead to severe joint destruction. Gout is the most frequent inflammatory arthritis in adult, affecting between 0.1 to 6.8% of the world population(1). This disease is particularly disabling, accounting for 1.3 million years lost due to disability in 2017(2).

Despite existing guidelines on management of the disease and widely available treatment of acute flares or chronic gouty arthritis(3,4), gout remains alarmingly undertreated. A recent global epidemiology study reports that only 30% to 50% of patients receive urate-lowering therapy and fewer than half of them adhere to treatment(1).

This gap highlights an urgent need to understand and address the reasons behind this undertreatment and the associated risk factors.

Despite the high prevalence of this disease, existing gout registers predominantly come from rheumatology settings(5,6) which do not fully capture the disease’s spectrum(7). One of the barriers to create larger gout registers are their labour-intensive nature, requiring manual data collection by healthcare professionals. Furthermore, administrative datasets, a common source for research studies, often lack relevant clinical information such as laboratory data and specific patient-centred outcomes(8). Notably inflexible, these datasets cannot be tailored to answer specific research questions(9). Built for insurance claim purpose, their ability to detect an actual patient illness can be low depending on the condition(10), and this is especially true for gout patient(11,12). Building registers directly from the electronic health records (EHR), a process proved feasible for chronic kidney disease(8), can help solve these shortcomings, streamline the process, and obtain a more complete picture of gout patients.

The aims of this article are to prove the feasibility of setting a reproducible gout register based on hospital EHR data, present the validity of its diagnostic algorithm, detail its implementation, and provide an overview of the resulting register for both out- and inpatients. The final objective is to enhance gout management and patient outcomes through a comprehensive and validated register, offering a deeper insight into the disease’s diagnosis and management.

## Methods

### Study setting

The Geneva University Hospitals (HUG) is a 2’000 beds French-speaking tertiary hospital, constituted of 8 hospital sites and 2 clinics. Every year, it cares for 60’000 inpatients, provides 1.2 million outpatient visits and receives close to 250’000 emergency room visits(13). Beyond providing the standard array of care for both inpatient and outpatient, it offers specialised care to psychiatric patients, inmates, and vulnerable populations.

### Health and administrative data sources

A common electronic health record is used in every hospital, clinic and various points of care belonging to the HUG. A dedicated software contains all administrative and medical information, where any health professional can retrieve and add data transversally. These data are stored on a centralised repository and mirrored in a MongoDB database.

The HUG laboratory is accredited for joint aspiration analysis by the Swiss Accreditation Service (norm 15189), and its technicians trained by accredited organizations, holding certificates of expertise in crystal evaluation.

### Inclusion criteria and time frame

All adults ≥ 18 years old, currently deceased or living, with any contact as an in- or outpatient with the Geneva University Hospital from January 1^st^ 2013 to the 31 of December 2022 were included in the queries to develop the register. The year 2013 was chosen because the Swiss diagnosis-related group (DRG) system was implemented in 2012. This system, used for insurance claim purpose in the inpatient setting in Switzerland, classifies patients and their diagnoses according to certain groups, which are similar in medical and economical term(14). The german-modification of the International Classification of Disease (ICD-10 GM) diagnostic codes play a preponderant role in this system.

### Criteria for potential gout cases

We assessed six criteria to capture gout diagnosis (table 1; for full detail see supplementary table 1).

**Table 1:** Criteria considered for gout diagnosis and their conditions. Gout-related terms included “gout”, “podagra”, “tophus”, “tophi”, “tophaceous” for the document, problem list and imaging report criteria. The latter included also “double-contour.

| Criteria considered for gout diagnosis | Conditions |
| --- | --- |
| 1. ICD-10-GM diagnosis code | M10.00-M10.99 |
| 2. Problem list of the EHR | Regular expression query for CPPD-related terms |
| 3. Joint aspiration result | Presence of monosodium urate crystals |
| 4. Medication | Allopurinol, Febuxostat, Probenecid or Lesinurad |
| 5. Documents (any reports) | Regular expression query for gout-related terms |
| 6. Imaging reports | Regular expression query for gout-related terms |

### Refining criteria for accurate diagnosis of gout

In a first step, we selected small groups of patients to verify and refine our criteria. Sample size for these initial queries was calculated based on expected positive predictive value (PPV) and tolerating a 5% half-confidence interval. For instance, for ICD codes, assuming a 95% PPV, and accepting a confidence interval between 90% and 100%, the computed sample size was 20 patients. The charts of these 20 patients were reviewed to refine the appropriate criteria. All M11 ICD-10-GM codes (i.e. other crystal arthropathies) were excluded for the ICD10 code criteria because most results were related to calcium pyrophosphate deposition disease.

For free-text searches (problem list, medical documents, and imaging reports) a list of proverbs, of medication and of human body liquids was built to detect false positives.

Indeed, ‘gout’, in French ‘goutte’, is a very common word, that can be used also for medication (drops), as a symptom in uro-gynecology (as blood or urine drop), in psychiatry (a proverb ‘the drop that made the vase overflow’, similar to ‘the straw that broke the camel back’), or even as surname. For free-text searches (problem list, medical documents, and imaging reports), presence of negation or double negation was examined in sentence related to gout, to identify situations where the text expressed an exclusion of gout diagnosis. The code and the different steps of the context analysis are described in supplementary material and the associated code is available at https://gitlab.unige.ch/goutte/register_validation. Allopurinol was often used in the oncology setting without a gout diagnosis. By excluding patients with an ICD-10-GM codes for leukaemia or lymphoma(15), we were able to exclude cases of allopurinol used for an oncologic indication and keep cases related to gout only.

### Diagnostic algorithm

Based on the previous criteria, we used the following algorithm to identify patients with gout, as any of the following 3 conditions:

1. A gout diagnosis in the problem list **OR**
2. Positive joint aspiration result for uric acid crystal **OR**
3. Any combination of at least **two** other criteria of the following variables:
  a. Medications
  b. ICD-10-GM codes
  c. Text of medical documents
  d. Text of joint imaging reports

### Sensitivity analysis

To test how the chosen diagnostic algorithm influenced the PPV and NPV of the register, we considered two alternative algorithms, one more sensitive and one more stringent than our main algorithm:

- Algorithm 1 (more sensitive): any of the 6 criteria
- Algorithm 2 (more stringent):
  - A gout diagnosis in the problem list **OR**
  - Positive joint aspiration result for uric acid crystal **OR**
  - Any combination of at least **three** of the remaining criteria

### Gout “gold standard” definition

To evaluate the accuracy of the queries to detect a real gout diagnosis and further optimize them, a randomly selected sample of charts were manually assessed. Every chart was reviewed by a physician and a research nurse. Disagreements were adjudicated by a rheumatologist. A diagnosis of gout was confirmed if documented by a physician in the patient’s medical records. Any text referring to a gout or a gout-related terms (i.e. tophi, podagra) was considered. If multiple differential diagnosis were mentioned, the final diagnosis established was considered. In case of a rheumatologic evaluation, the diagnosis of the rheumatologist had priority.

Patients with a history of gout, without any feature of gout during any episode of medical care, were considered as having gout if established as such by a doctor in the charts.

Mono or oligo-arthritis with feature of gout (rapid onset of pain, response to colchicine, NSAID or corticoid, and no other apparent cause) but without a specified diagnosis by the team in charge, were classified as equivocal. Use of a urate lowering therapy without any documented gout diagnoses were also considered equivocal.

### Sample size calculation

For PPV, assuming a 95% PPV, with a precision of +-2% (93% to 97% confidence interval – CI), the calculated sample size corresponded to at least 456 patients. We applied the same conditions for NPV, yielding the same minimal number of patients. To respect the proportions of patients for each query and to account for potential incomplete information in EHR, 518 charts were extracted for the positive predictive value and 492 for the negative predictive value assessment.

### Selection of non-gout cases at risk of developing gout

To calculate the negative predictive value of the criteria, patients at risk of developing gout but not detected as having gout by our algorithm were selected, based on a combination of known risk factors(1,16,17).

1. Sex ≥ 65 for women and ≥ 40 for men **AND**
2. Overweight or obesity (Body-mass index > 25kg/m^2^) **AND**
3. Any of the following:
  a. Metabolic syndrome
  b. Myocardial infarction
  c. Chronic kidney disease
  d. Deleterious use of alcohol
  e. Hyperuricaemia (>500 μmol/l or 8.4 mg/dl)

The detail of the criteria used can be found in the Supplementary table 2.

### In- and outpatient differentiation

We categorised patients as in- or outpatients according to the setting where they first met the condition to be diagnosed by our algorithm. For example, a patient with a positive aspiration in the ambulatory setting was categorised as outpatient, even if they later received an ICD-10 GM code in an inpatient setting. If two criteria were required, the setting where the second criterion was met determined the patient’s classification as either inpatient or outpatient.

### Additional variables

In addition to the criterion necessary to determine whether a patient had gout and dates on which each of the criterion were recorded, relevant information was automatically collected. They included anthropometric & demographic data, information regarding gout episodes, clinical pathway, comorbidities, laboratory values, joint aspirations, drugs, and presence of a rheumatology consultation (Table 4 & Supplementary table 3).

Patients were also classified as having a high gout burden if they experienced a gout episode that required hospitalization (primary diagnosis of gout) or had at least two separate gout flares occurring at least 30 days apart but within a one-year period, as confirmed by joint aspiration.

### Statistical analysis

We summarised data using frequencies and percentages for categorical variables and means and standard deviations for continuous variables. Positive predictive value was calculated as the proportion of gout patients according to chart review divided by the total number selected for review. Equivocal were considered as not having gout.

Negative predictive value was calculated as the proportion of patients not having gout according to chart review divided by the total number selected for review. Equivocal were considered as having gout.

Confidence intervals were computed using the Agresti and Coull method(18). All statistics were computed using the software R V4.3.0(19).

### Ethical consideration

The creation and use of the register for quality improvement programs has been approved by the Geneva ethics commission (CCER 2023-00129).

## Results

### Process leading to the register

A total of 2,110,902 unique patients were seen at the hospital over 10 years, of which 10,289 had at least one criterion for gout. Of these, 5,151 were detected only by a single criterion other than the problem list and the joint aspiration (i.e., only ULT, document, imaging report or ICD-10-GM), and were excluded from the register by our main diagnostic algorithm, yielding a final register of 5,138 gout patients (figure 1). This corresponds to an incidence of 2.4 diagnoses per year per 1,000 patients.

### Criteria combination

Amongst all patients selected by at least one criterion (figure 2A), most were detected from documents alone (28.4%), by use of a drug prescription alone (18.9%), or by a combination of both (7.26%). It was then followed by the combination of the problem list and the presence of a diagnostic in a document (5.7%). When selecting patients based on our diagnostic algorithm (Problem list OR Joint aspiration OR ≥ 2 Other criteria, figure 2B), the vast majority of the 5,151 rejected patients were those detected from documents alone or by a drug prescription alone. The combination documents/drugs (14.5%) and problem list/documents (11.5%) were then the most frequent.

Interestingly, 2.0% of patients had a positive aspiration for uric acid crystal without any gout diagnosis. Outpatients and inpatients showed relatively similar patterns of criteria presentation (Supplementary figure 1A and 1B), though a positive aspiration without further documentation was slightly more frequent for outpatients (3.0%) than for inpatients (1.7%).

### Positive predictive value

Our diagnostic algorithm led to a PPV of 92.4% (95%CI: 88.5 to 95.0%, see table 2). Results were similar in the in- and outpatient setting (93.3% and 92.4% respectively, see Supplementary tables 6 and 7).

**Table 2:**
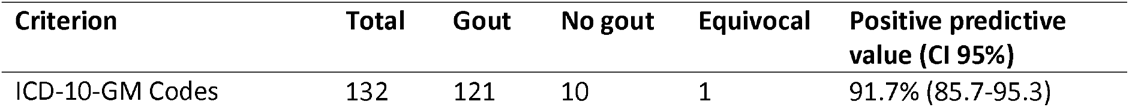
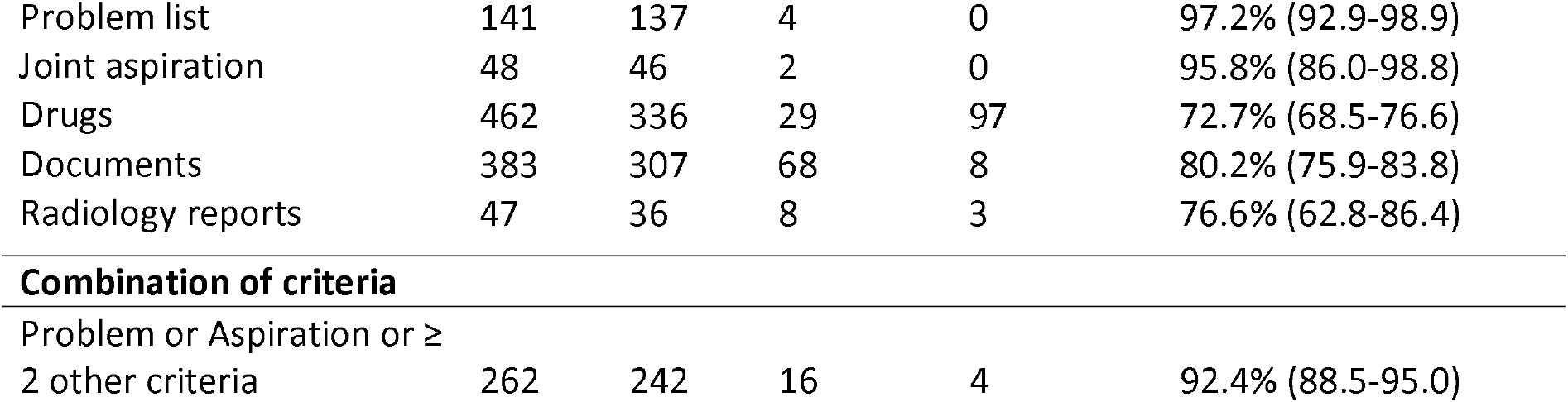
Presence of a written gout diagnosis (defined gold standard) for each criterion and associated positive predicted value (PPV) (a patient can appear in multiple criteria) amongst 518 manually reviewed charts. PPV was calculated as the number of gout patients divided by the total number of patients, meaning that equivocal cases were classified as non-gout in the PPV analysis.

Positive predictive value
| Criterion | Total | Gout | No gout | Equivocal | Positive predictive value (CI 95%) |
| --- | --- | --- | --- | --- | --- |
| ICD-10-GM Codes | 132 | 121 | 10 | 1 | 91.7% (85.7-95.3) |
| Problem list | 141 | 137 | 4 | 0 | 97.2% (92.9-98.9) |
| Joint aspiration | 48 | 46 | 2 | 0 | 95.8% (86.0-98.8) |
| Drugs | 462 | 336 | 29 | 97 | 72.7% (68.5-76.6) |
| Documents | 383 | 307 | 68 | 8 | 80.2% (75.9-83.8) |
| Radiology reports | 47 | 36 | 8 | 3 | 76.6% (62.8-86.4) |

*Table 2: Presence of a written gout diagnosis (defined gold standard) for each criterion and associated positive predicted value (PPV) (a patient can appear in multiple criteria) amongst 518 manually reviewed charts.
|  |  |  |  |  |  |
| --- | --- | --- | --- | --- | --- |
| Problem or Aspiration or ≥ 2 other criteria | 262 | 242 | 16 | 4 | 92.4% (88.5-95.0) |

Individual criteria within our algorithm exhibited variable PPVs. The highest values were obtained for problem list and joint aspiration (97.2% (95%CI: 92.9 to 98.9) and 95.8% (95%CI: 86.0 to 98.8) respectively). In contrast, the PPVs were lower for drug (72.7% (95%CI: 68.5 to 76.6)), document (80.2% % (95%CI: 75.9 to 83.8)) and radiological reports (76.6% (95%CI: 62.8 to 86.4)). Notably, most patients without gout identified by these criteria were exclusively detected by a single criterion, for which ICD-10, drugs, documents, and radiological reports yielded suboptimal VPP (Supplementary table 5).

### Negative predictive value

Amongst the 2,110,902 patients seen at the hospital over 10 years, 15,646 had at least one risk factor for gout (Supplementary table 8). Of these, 2,588 (16.5%) were found to have a gout by our diagnostic algorithm, and were excluded of this sample to estimate NPV, yielding 13,058 patients with risk factors.

Amongst these highly at-risk patients, NPV for all risk factors (Table 3) was excellent, except for the uricaemia criterion. The overall NPV was 94.3% (95%CI: 91.9 to 96.0). Patients with a single risk factor (i.e. without any other NPV criterion) yielded similar results (Supplementary table 9).

**Table 3:** Presence of a written gout diagnosis (defined gold standard) amongst patients with risk factors for gout but not detected by the algorithm and associated negative predictive value (NPV) (a patient can appear in multiple risk factors) from 492 manually reviewed charts. NPV was calculated by dividing the number of patients not having gout by the total number of patients, meaning that equivocal cases were classified as gout in NPV analysis.

| Criterion | Total | Gout | No gout | Equivocal | Negative predictive value (CI 95%) |
| --- | --- | --- | --- | --- | --- |
| **Age ≥ 65 for women OR ≥ 40 for men AND Overweight (Body-mass index > 25kg/m<sup>2</sup>) AND:** |  |  |  |  |  |
| Uricaemia > 500 µmol/l | 41 | 8 | 33 | 0 | 80.5% (66.0 to 89.8) |
| Chronic kidney disease | 154 | 10 | 142 | 2 | 92.2% (86.9 to 95.5) |
| Metabolic syndrome | 79 | 6 | 73 | 0 | 92.4% (84.4 to 96.5) |
| Myocardial infarction | 196 | 2 | 193 | 1 | 98.5% (95.6 to 99.5) |
| Deleterious use of alcohol | 136 | 3 | 132 | 1 | 97.1% (92.7 to 98.9) |
| **Total (any risk factor)** | **492** | **24** | **464** | **4** | **94.3% (91.9 to 96.0)** |

### Sensitivity analysis

The more sensitive algorithm, considering any criterion as enough to diagnose patients with gout, would have included all 10,289 patients, with a PPV of 69.1% (95%CI: 65.0 to 72.9%), and an NPV of 97.5% (95%CI: 95.6 to 98.6).

The more stringent algorithm would have included only 3746 patients, with a PPV of 96.9% (95%CI: 93.5 to 98.6%) and an NPV of 91.6% (95%CI: 88.8 to 93.7%).

### Gout register

The 5,138 patients detected by our algorithm were mostly old men, frequently overweight (Table 4). The vast majority had at least one comorbidity (83%), hypertension (69.4%), cardiovascular and ischaemic diseases (stroke, heart failure, ischaemic heart and peripheral vessel disease, 53%) being the most prevalent. As of 31.12.2022, 34.5% of the patients were recorded as deceased. Most patients were Swiss citizen (68.8 %) or came from other European countries (22.6 %). At time of detection by the algorithm, 74.3% were categorised as inpatients. Patients were mostly detected in the department of medicine (27.8%), followed by geriatrics (19.5%). 92% of the patients had a document referring the gout diagnosis, and 18.6% had a joint aspiration positive for monosodium uric acid crystal. A gout diagnosis was documented in the problem list in half of the case overall (53.3%), but reached 78.2% in the outpatient setting. Around half the patients (49.2%) had an ICD10 code corresponding to gout. 6.7% of the patients had a gout that led to a hospitalisation or at least two flares within a year. Concerning drugs, 57.0% of the patients had received an ULT at any one time, most frequently allopurinol, and 48.3% received colchicine. Uricosurics (probenecid and lesinurad) were almost never prescribed. Only a third of the patients (33.0%) had a rheumatology consultation.

**Table 4:** Characteristics and stratification per setting of care of the patients forming the final register. BMI: body-mass index. ^§^High gout burden is defined as patients that had a gout leading to a hospitalisation or ≥ 2 flares proven by joint aspiration occurring at least 30 days apart but within a one-year period. ^†^Details of the ICD-10 codes used to assess comorbidities can be found in supplementary table 3. There are no missing value for socio-demographic variables, except for the BMI (27.1%).

| Setting \ Variables | Overall | Outpatient | Inpatient |
| --- | --- | --- | --- |
| Patient in the register | 5138 | 1320 | 3818 |
| Total number of patients | 2110902 | 1806981 | 860049 |
| Incidence (per 1000-person year) | 2.4 | 0.7 | 4.4 |
| Age (mean (SD)) | 71.22 (14.89) | 66.25 (14.88) | 72.93 (14.50) |
| Men (%) | 3931 (76.5) | 1062 (80.5) | 2869 (75.1) |
| BMI (mean (SD)) | 28.33 (6.54) | 28.47 (6.06) | 28.28 (6.71) |
| BMI 25-30 kg/m<sup>2</sup> | 1381 (36.9) | 374 (37.0) | 1007 (36.8) |
| BMI ≥ 30 kg/m<sup>2</sup> | 1220 (32.6) | 351 (35.1%) | 869 (31.8%) |
| Number of death (%) at 31.12.2022 | 1771 (34.5) | 286 (21.7) | 1485 (38.9) |
| Criterion ever present |  |  |  |
| ICD-10-GM Codes | 2528 (49.2) | 485 (36.7) | 2043 (53.5) |
| Problem list | 2741 (53.3) | 1032 (78.2) | 1709 (44.8) |
| Joint aspiration | 957 (18.6) | 236 (17.9) | 721 (18.9) |
| Drugs | 2807 (54.6) | 663 (50.2) | 2144 (56.2) |
| Documents | 4728 (92.0) | 1157 (87.7) | 3571 (93.5) |
| Radiology reports | 698 (13.6) | 182 (13.8) | 516 (13.5) |
| Urate lowering therapy (ever) |  |  |  |
| Allopurinol | 2813 (54.7) | 695 (52.7) | 2118 (55.5) |
| Febuxostat | 209 ( 4.1) | 64 ( 4.8) | 145 ( 3.8) |
| Probenecid | 9 ( 0.2) | 6 ( 0.5) | 3 ( 0.1) |
| Lesinurad | 1 ( 0.0) | 1 ( 0.1) | 0 ( 0.0) |
| None | 2189 (42.6) | 592 (44.8) | 1597 (41.8) |
| Colchicine (ever) | 2484 (48.3) | 602 (45.6) | 1882 (49.3) |
| Rheumatology consultation (ever) | 1690 (32.9) | 376 (28.5) | 1314 (34.4) |
| High gout burden<sup>§</sup> | 344 ( 6.7) | 79 ( 6.0) | 265 ( 6.9) |
| ≥2 flare / year | 76 ( 1.5) | 26 ( 2.0) | 50 ( 1.3) |
| Hospitalisation due to gout | 294 ( 5.7) | 63 ( 4.8) | 231 ( 6.1) |
| Comorbidities (ICD-10-GM codes)<sup>†</sup> |  |  |  |
| Number (median [IQR]) | 3 [1, 5] | 3 [1, 5] | 3 [2, 5] |
| Any comorbidity | 4266 (83.0) | 997 (75.5) | 3269 (85.6) |
| Hypertension | 3564 (69.4) | 814 (61.7) | 2750 (72.0) |
| Dyslipidaemia | 1822 (35.5) | 453 (34.3) | 1369 (35.9) |
| Diabetes | 1524 (29.7) | 355 (26.9) | 1169 (30.6) |
| Cardiovascular diseases | 2722 (53.0) | 566 (42.9) | 2156 (56.5) |
| Liver disease | 603 (11.7) | 149 (11.3) | 454 (11.9) |
| Kidney disease (Stage ≥ 3) | 1818 (35.4) | 403 (30.5) | 1415 (37.1) |
| Psychiatric disorder | 1983 (38.6) | 448 (33.9) | 1535 (40.2) |
| Alcohol use disorder | 733 (14.3) | 172 (13.0) | 561 (14.7) |
| Organ transplant | 84 ( 1.6) | 33 ( 2.5) | 51 ( 1.3) |
| Malignancies | 1145 (22.3) | 306 (23.2) | 839 (22.0) |
| Purine disease | 107 ( 2.1) | 22 ( 1.7) | 85 ( 2.2) |
| Department at first algorithm detection |  |  |  |
| Medicine | 1428 (27.8) | 544 (41.2) | 884 (23.2) |
| Geriatrics | 1002 (19.5) | 46 ( 3.5) | 956 (25.0) |
| Surgery | 718 (14.0) | 264 (20.0) | 454 (11.9) |
| Acute medicine | 681 (13.3) | 32 ( 2.4) | 649 (17.0) |
| Primary care | 223 ( 4.3) | 111 ( 8.4) | 112 ( 2.9) |
| Psychiatry | 53 ( 1.0) | 20 ( 1.5) | 33 ( 0.9) |
| Other | 1033 (20.1) | 303 (23.0) | 730 (19.1) |

Our algorithm revealed a 30% decrease in yearly gout diagnoses, falling from 2.9 to less than 2 per 1,000 patient-years before and since the COVID-19 pandemic in 2020 (figure 3). This decline remained consistent over time and was primarily observed in the inpatient setting (Supplementary figure 2).

When studying how our algorithm’s criteria have evolved over time to initially diagnose patients with gout (second detection in the case of combination of criteria, Supplementary figure 2), we observed a rise in the problem list over time, introduced gradually in our hospital since 2011. Notably, there was a decreasing trend in joint aspiration as the initial detection method, particularly in inpatient settings since the onset of the COVID-19 pandemic in 2020.

## Discussion

This study demonstrates the feasibility and the relevance of building a clinical gout register through automated queries on EHR data, encompassing out- and inpatients across diverse care settings. Out of over 2 million patients, 5,138 were definitively diagnosed with gout, reflecting an incidence rate of 2.4 per 1,000 patients annually.

The fine tuning of our criteria on a small subset of patients together with the careful estimation of the validity of various algorithms allowed us to propose an efficient algorithm with excellent positive and negative predictive values, ensuring accurate identification of gout patients. The incidence of newly diagnosed patients up to 2019 (pre-COVID-19) is comparable to previously developed medical records databases across the world(1). It is twice that reported in studies exclusively relying on ICD-10 codes(20). This aligns with our finding that over half of our diagnosed patients did not have ICD-10 codes for gout. Our approach based on a combination of various criteria offers thus a less biased mean to study this disease and underscores the value of using a multifaceted approach.

The decline in gout diagnoses since the onset of the COVID-19 pandemic, the limited use of urate-lowering therapy or the low rate of rheumatology consultation show that our approach offers insights into diverse aspects of patient care.

The yearly incidence of patients diagnosed by our algorithm decreased from 2020 onwards, corresponding to the beginning of the COVID-19 pandemic, in line with finding from a recent study in England(20). This decrease could be the consequence of 3 different factors: the decrease of diagnosis by healthcare professionals, the change of population attending the hospital, or an actual decrease of the gout incidence. First, since the start of the pandemics, the disruption of medical education of physicians and medical students could have affected their ability to detect and diagnose gout(21,22). Second, the lack of access to healthcare for gout patients due to the COVID-19 pandemics, with unrecognition of the disease and inability to refill prescription drug could explain the inflexion of diagnoses (23), with potential lasting effect(20). Third, although it seems unlikely that SARS-CoV-2 affected directly the occurrence of gout, it imposed a great toll on patient with cardiovascular risk factors(24,25). This population of patient, particularly prone to gout, could have been reduced.

In our register, the best criterion (highest PPV) to detect a gout diagnosis was the problem list, which was gradually introduced since 2011 and is now used in every department. Problem lists keep track of all current and past diagnoses, they centralise the usually scattered relevant medical information’s and are used to familiarise oneself with a new patient(26). In 2022, it was a prevalent inclusion criterion in the register for both out- and inpatient. The outpatient setting saw higher prevalence, as it was introduced earlier in our EHR. It is, however, as a manually created and edited tool, prone to inaccuracies, accumulation of duplicate, lack of update and incompleteness(27). Efforts to maintain their quality are warranted.

Despite being the gold standard, it is noteworthy that the presence of acid uric crystals in the synovial fluid did not yield a 100% PPV. Indeed there can be a lack of consensus between operators in analysing synovial fluid, even in an accredited laboratory resulting in false positive uric acid crystal results(28) 29).

Despite its rather good PPV, in agreement with what has been reported in the literature(12,30), the use of ICD codes alone was not sufficient to build a register of gout patients. Indeed, half of the patients identified did not have an ICD gout diagnosis, either because they were never hospitalised, wrongly coded or not coded at all. Studies have shown mixed result for the use of ICD codes as predictor of a gout diagnosis(10,12,30). In the Swiss healthcare system, ICD codes are documented by specialised coders, based on a written diagnosis in the EHR, either as a problem list or in the final report. This could lead to under reporting of the disease and lack of proper billing.

The estimated PPV of the drug criteria was not optimal partly due to the lack of gout diagnosis by a clinician in the EHR (our defined gold standard). Many patients had already been prescribed ULT outside this hospital, probably due to a history of gout. However, ULT might have been prescribed for other reasons such as kidney stone without gout, oncological indication, or inappropriately for asymptomatic hyperuricaemia. In our study, most charts did not provide other diagnosis explaining the need for a urate lowering therapy. Indication alert prompting the documentation of gout in the problem list, triggered by the prescription of an urate lowering therapy, could help solve this shortcoming(31).

The query in documents was complicated by the fact that gout, a prevalent word in French, is used commonly by patients and healthcare professionals. We used a combination of regular expression and natural language processing to exclude situations where the word gout was used in a negative context (e.g. psychiatry) or referring to drugs (e.g., drops of vitamins) and analyses (e.g., drops of blood). Although some researchers have proposed artificial-intelligence-based models to extract data from EHR with great success(32,33), queries-based algorithms like ours have also succeeded with the advantage of simplicity and easy reproducibility(34). Use of advanced natural language processing or large language model could help find the correct diagnosis when multiple differential diagnoses are mentioned, though preliminary efforts in our hospital yielded lower predictive values. Indeed, despite best optimisation of the regular expression filters, during an episode of acute arthritis, gout was often considered and mentioned in clinical documentation, especially admission record, before being discarded as the final diagnosis.

There are several strengths of our study. First, we provided detailed performance metrics of our diagnosis algorithm, using conservative choices to ensure accuracy. Indeed, we used an at-risk population for the negative predictive value, and equivocal cases were considered as not having gout in the PPV or as having gout in the NPV calculation. Second, we proposed sensitivity analysis regarding two alternative diagnosis algorithms. Third, by providing a detailed procedure and choosing commonly documented variables we facilitate the implementation of similar registers in other hospitals. Last, our approach uniquely sets apart this register from others registers, such as the CORRONA and EHR-based RISE register which are confined to rheumatology practices(5,6). We included a very diverse population of out- and inpatients, from all specialties of an academic tertiary hospital, including vulnerable population (uninsured, migrants, inmate, …), thereby providing a more comprehensive and varied dataset than typically seen in specialty-specific registers.

The main limitation of this register is selection bias. As in all hospital EHR-based study (e.g., chronic kidney disease register(8)), this register only contains patients who used medical resources in the ecosystem of the Geneva university hospital. It does not assess patients consulting only in private clinics or practices, nor those who did not seek medical attention. This risk is mitigated by the fact that the Geneva University Hospitals is the only public hospital in the region, providing free in- and outpatients care to vulnerable population and inpatient care for the majority of the regional population. This is further confirmed by the high incidence of new cases reported by our method. Another limitation is the use of a single hospital for the register, due to the use of different EHR systems in Switzerland. Finally, the gout diagnosis by a physician in the hospital can be seen more as a silver standard, since patients could have been diagnosed outside of the hospital, which may certainly bias prevalence studies.

This study proves the feasibility of implementing an electronic health-record based register with excellent positive and negative predictive values for detecting gout patient. The use of criteria based on several variables allowed to detect gout diagnosis otherwise missed by ICD codes or explicit diagnosis alone. The automatic nature of the query makes this register inexpensive and sustainable, facilitating the assessment of the adequacy of gout management, the monitoring of indicators following quality improvement projects, and the detection of gout patients to be included in new studies or trials. Also the decline of gout diagnoses since 2020, especially evident in inpatient settings, prompts questions about how the pandemic may have affected healthcare access, patient behaviours, and diagnostic approaches.

## Data Availability

All data produced in the present study are available upon reasonable request to the authors

## Acknowledgment

We thank Dr Anne-Marie Rassinoux and Mr Emmanuel Durand from the direction of the information system of the HUG for providing access to the data lake containing the EHR datas.

A feasibility study of the creation of the register was presented at the EULAR 2023 conference(35).

## Contributorship

Nils Bürgisser and Denis Mongin contributed equally to this study.

Designed research: KL, DSC

Performed research: NB, DM, SM

Analysed Data: DM, NB

Wrote the Paper: NB, DM, KL, DSC

Critical revision of the paper: all authors

## Figure legends

*Figure 1. Flow chart of the patient selection process leading to the final gout register. ULT: urate-lowering therapy. ICD-10-GM: german-modification of the International Classification of Disease, 10 ^th^ revision*

*Figure 2A and B: Upset-plot of the six criteria identifying gout patients in the electronic health record of the Geneva University Hospital. Figure 2A depicts the combinations of criteria present amongst patients selected by at least one criteria (n=10,289). Figure 2B depicts the combinations of criteria present amongst patients selected by the final algorithm used for the register (Problem list OR Aspiration OR ≥ 2 other criteria). Rare combinations of criteria are not displayed. Stratification by setting (inpatient/outpatient) can be found in supplementary figure 1.*

*Figure 3: Evolution in time of number of gout patient per year (left y axis) and corresponding incidence (red line, right y axis)*

