## Supplementary material for "Enhancing gout management by creating a register using automated queries in electronic health records"

| Criteria considered for gout diagnosis | Included for gout diagnosis | Excluded following exploratory phase |
|---|---|---|
| **1. ICD-10-GM diagnosis code** | M10.00-M10.99: gout | M11.X: other crystal arthropathies. |
| **2. Documents**<br>• Inpatient report (Admission or discharge letter from any type of care)<br>• Outpatient report (emergency room, specialist consultation) | Regular expression query in full text for:<br>• “gout”, “podagra”, “tophus”, “tophi”, “tophaceous”[†] | Exclusion of documents using a defined set of regular expression[§] |
| **3. Problem list of the EHR** (independent of the ICD-10-GM code) | Presence of one of the following keywords:<br>• “gout”, “gouty”, “podagra”, “tophus”, “tophi”, “tophaceous”[†] | Exclusion of a set list of words<br>See supplementary table 3 |
| **4. Joint aspiration result** | Presence of monosodium urate crystals | None |
| **5. Medication**<br>• Prescription of urate lowering therapy (ULT)<br>• Prescription form containing an ULT | At least one of:<br>• Allopurinol (ATC: M04AA01)<br>• Febuxostat (ATC: M04AA03)<br>• Probenecid (ATC: M04AB01)<br>• Lesinurad (ATC: M04AB05) | Prescription of an ULT and presence of at least one of the following ICD-10 GM code at any time:<br>C81 to C96: Malignant neoplasms, stated or presumed to be primary, of lymphoid, haematopoietic and related tissue |
| **6. Imaging report**<br>• X-ray<br>• ultrasound<br>• dual-energy computed tomogram | Regular expression query for the following words:<br>• “gout”, “podagra”, “tophus”, “tophi”, “tophaceous”, “double contour”[†] | Exclusion of documents using a defined set of regular expression [§] |

*Supplementary table 1 : Variables and criterion used to assess gout diagnosis. *Benzbromarone isn’t authorized on the Swiss market. Lesinurad was pull out of the market in 2021. Rasburicase isn’t authorized for chronic gout treatment. † French translation of the original words used are described in the supplementary table 3. § the R code, as well as a working example has been made available here: https://gitlab.unige.ch/goutte/register_validation. ICD-10-GM: German Modifications of the International Classification of Diseases, 10th revision ATC: Anatomical Therapeutic Chemical Classification*

| Combination | | Variables considered for risk factors | Criterion for gout risk factors (and ICD-10 GM codes) |
|---|---|---|---|
| **AND** | | **Sex and age** | **Women ≥ 65 years old**<br>**Men ≥ 40 years old** |
| **AND** | | **BMI** | **> 25 kg/m$^2$** |
| **AND** | OR | Metabolic syndrome | • Diabetes or glucose intolerance (E10.X to E14.X, R73.X)<br>• Hypertension (I10.X to I15.X)<br>• Dyslipidaemia (E78.x)<br>• BMI > 30 kg/m$^2$ |
| | OR | Myocardial infarction | I21.X to I25.X |
| | OR | Chronic kidney disease | N18.X<br>Problem list: Chronic kidney disease G3a or worse (including dialyses) |
| | OR | Lifestyle at risk to develop a gout | Deleterious use of alcohol (F10.1, F10.2, F10.3) |
| | OR | Uricaemia | Serum urate > 500 µmol/l (8.4 mg/dl) |

*Supplementary table 2: Variables and criteria used to assess gout risk factors.*

| Category | Variables |
|---|---|
| Anthropometric & demographic data | Age<br>Sex<br>Weight<br>Height<br>BMI<br>First language<br>Country of residency<br>Nationality<br>Insurance class<br>Living/death status<br>Marital status |
| Clinical pathway | Inpatient gout episode of care<br>Outpatient gout episode of care<br>Departments involved<br>Duration of hospitalisation |
| Diagnosis | Main diagnosis (inpatient) |
| Comorbidities (ICD-10-GM diagnostic) | Number of comorbidities<br>Individual comorbidities<br>Hypertension and hypertensive diseases:<br>• Essential hypertension: I10.00 – I10.91<br>• Hypertensive cardiopathy: I11.00 – I11.91<br>• Hypertensive nephropathy: I12.00 – I12.91<br>• Hypertensive cardionephropathy: I13.00-I13.91<br>• Secondary hypertension: I15.00 – I15.91<br>Dyslipidaemia:<br>• Disorders of lipoprotein metabolism and other lipidaemias: E78.0-E78.9<br>Diabetes<br>• Type 1 diabetes: E10.01-E10.91<br>• Type 2 diabetes: E11.01 – E11.91<br>• Malnutrition diabetes: E12.11 – E12.91<br>• Other specified diabetes mellitus: E13.01 – E13.91<br>• Unspecified diabetes mellitus: E14.01 – E14.91<br>Cardiovascular and ischaemic heart disease:<br>• Angina pectoris: I20.00-I20-9<br>• Acute myocardial infarction: I21.0 – I21.9<br>• Recurring myocardial infarction: I22.0-I22.9<br>• Complication of a myocardial infarction: I23.0 – I23.8<br>• Other ischaemic cardiopathy: I24.0-I24.9<br>• Chronic ischaemic cardiopathy: I25.0-I25.9<br>• Cerebral infarction: I63.0 – I63.9<br>• Stroke, not specified: I64 |

| | |
|---|---|
| | • Occlusion and stenosis of precerebral arteries, not resulting in cerebral infarction: I65.0 – I65.9<br>• Occlusion and stenosis of cerebral arteries, not resulting in cerebral infarction: I66.0-I66.9<br>• Sequelae of cerebrovascular disease: I69.0 – I69.8<br>• Atherosclerosis: I70.0 – I70.9<br>• Arterial embolism and thrombosis: I74.0 – I74.9<br>• Heart failure: I50.0 – I50.9<br>Chronic kidney disease (stage ≥3): N18.3 – N18.9<br>Diseases of liver: K70-K77.8<br>Psychiatric disorders: F00-F99<br>Transplanted organ and tissue status: Z94.0 – Z94.9<br>Malignancies: C00-C97<br>Disorders of purine and pyrimidine metabolism: E79.1-E79.9 |
| Laboratory data | Serum urate value<br>Serum creatinine and eGFR<br>CRP<br>Erythrocyte sedimentation rate<br>Liver function tests<br>Blood cell count |
| Joint puncture | Joint<br>Number of joint aspiration<br>Volume of synovial fluid<br>Presence of bacteriologic exam, Result of bacteriologic exam<br>Presence of cell count<br>Result of cell count<br>Presence of crystal evaluation<br>Crystal type in joint aspiration |
| Chronic drug treatment | Allopurinol<br>Febuxostat<br>Probenecid<br>Lesinurad<br>Rasburicase |
| Drug prophylaxis during initiation of ULT | Colchicine<br>Corticosteroid<br>NSAID |
| Acute flare treatment | Colchicine<br>Intra-articular corticoid injection (triamcinolone acetonide, betamethasone, triamcinolone hexacetonide)<br>Oral, IV or IM corticoid (dexamethasone, prednisone, methylprednisolone, prednisolone)<br>Anakinra or canakinumab<br>NSAID |
| Gout predisposing drugs | Aspirin<br>Ciclosporin |

| | |
|---|---|
| | Tacrolimus<br>Loop diuretics (furosemid, torasemid)<br>Thiazide diuretics (hydrochlorothiazide)<br>Thiazide-like diuretics (metolazone, indapamide, chlorthalidone) |
| Gout protecting drugs | Fenofibrate<br>Losartan<br>SGLT-2 inhibitors |
| Pain score | Visual analog scale |
| Fall | Fall event during an acute gout flare<br>(“déclaration de chute”) |
| Rheumatology consultation | Presence or absence |
| Imaging | Presence of an X-ray of the painful articulation<br>Presence of a CT dual energy of the painful articulation<br>Presence of an ultrasound of the painful articulation<br>Gout-related damage or tophi or double contour sign |

*Supplementary Table 3: Data extracted from the electronic health record.*

| Query | French translation | English translation |
|---|---|---|
| Documents | • « goutte »<br>• « podagre »<br>• « tophus »<br>• « tophi »<br>• « tophacée » | • Gout<br>• Podagra<br>• Tophus<br>• Tophi<br>• Tophaceous |
| Search in the problem list of the EHR (independent of the ICD-10-GM code) | • « goutte »<br>• « goutteux »<br>• « goutteuse »<br>• « podagra »<br>• « tophus »<br>• « tophi »<br>• « tophacée » | • Gout<br>• Gouty<br>• Gouty<br>• Podagra<br>• Tophus<br>• Tophi<br>• Tophaceous |
| Imaging report | • « goutte »<br>• « podagre »<br>• « tophus »<br>• « tophi »<br>• « tophacée »<br>• « double-contour » | • Gout<br>• Podagra<br>• Tophus<br>• Tophi<br>• Tophaceous<br>• Double contour |

*Supplementary table 4: original translation of the words used in the queries to detect gout patients.*

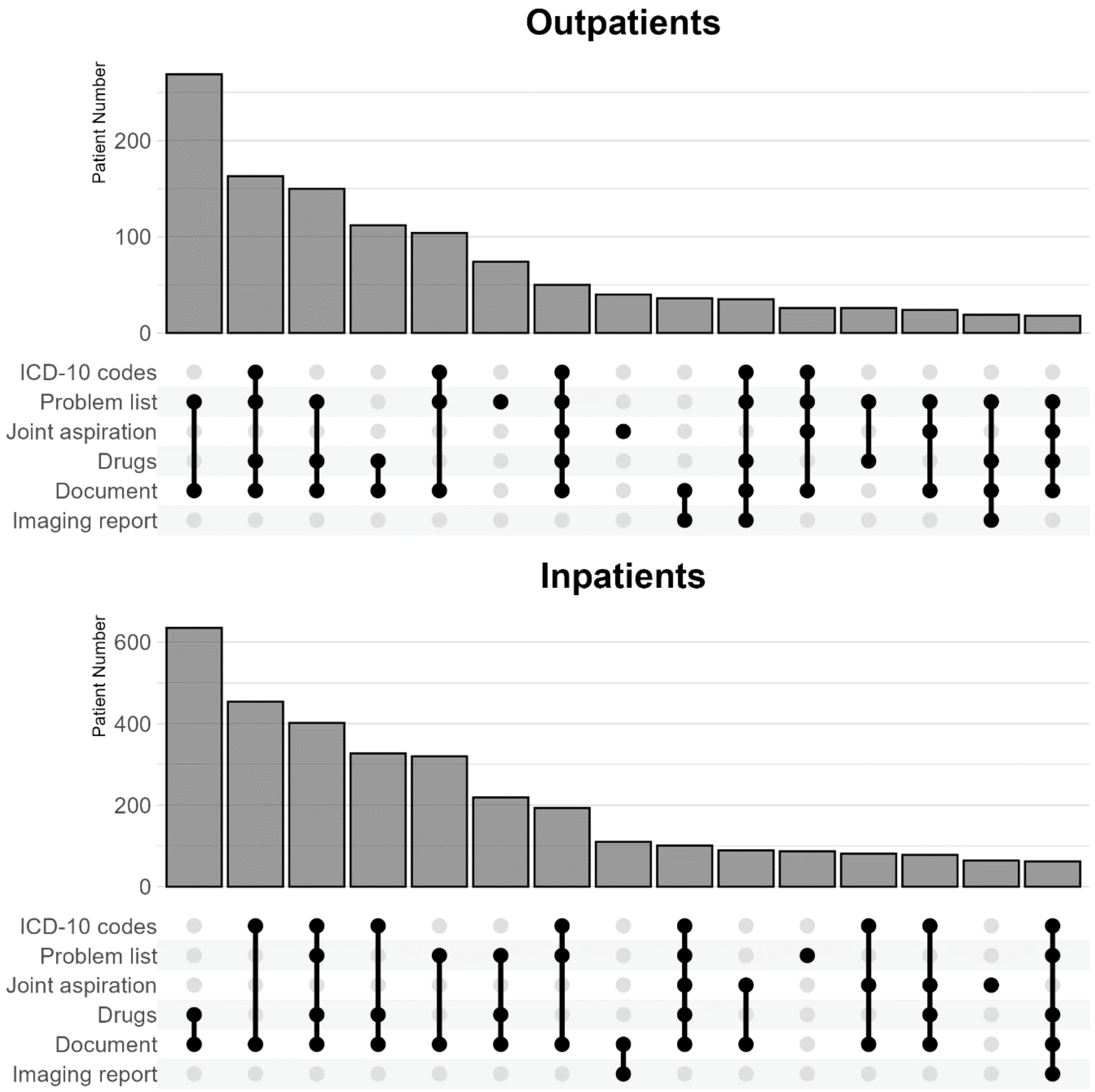


*Supplementary Figure 1A and 1B: Upset-plot of the six queries identifying gout patients in the electronic health record of the Geneva University Hospital when using a combination of criteria (Problem list OR Aspiration OR ≥ 2 other criteria) and by setting stratification. Rare combinations of criteria were not displayed. ICD-10-GM diagnoses are coded only during an inpatient stay*

| Criterion present alone | Total | Gout | No gout | Equivocal | Positive predictive value (CI 95%) |
|---|---|---|---|---|---|
| ICD-10-GM Codes | 5 | 2 | 3 | 0 | 40.0% (11.8-76.9) |
| Problem list | 8 | 7 | 1 | 0 | 87.5% (52.9-99.4) |
| Aspirations | 6 | 4 | 2 | 0 | 66.7% (30.0-90.3) |
| Drugs | 178 | 58 | 28 | 92 | 32.6% (26.1-39.8) |
| Documents | 143 | 84 | 55 | 4 | 58.7% (50.5-66.5) |
| Radiology reports | 9 | 2 | 5 | 2 | 22.2% (6.3-54.7) |

*Supplementary table 5 : Absolute numbers of patients and positive predictive value (PPV) of each criterion alone (a patient is detected only by the criterion). PPV was calculated by considering gout present versus gout absent or equivocal meaning equivocal cases were classified as non-gout in the PPV analysis.*

| Query (inpatients) | Total | Gout | No gout | Equivocal | Positive predictive value (CI 95%) |
|---|---|---|---|---|---|
| **Present irrespective of other criteria** | | | | | |
| ICD-10-GM Codes | 96 | 89 | 6 | 1 | 92.7% (85.7-96.4) |
| Problem list | 75 | 74 | 1 | 0 | 98.7% (92.8-99.9) |
| Joint aspiration | 38 | 37 | 1 | 0 | 97.4% (86.5-99.9) |
| Drugs | 208 | 202 | 1 | 5 | 97.1% (93.9-98.7) |
| Documents | 167 | 156 | 8 | 3 | 93.4% (88.6-96.3) |
| Radiology reports | 24 | 24 | 0 | 0 | 100.0% (86.2-100.0) |
| **Present alone** | | | | | |
| ICD-10-GM Codes | 0 | 0 | 0 | 0 | |
| Problem list | 4 | 4 | 0 | 0 | 100.0% (51.0-100.0) |
| Joint aspiration | 5 | 4 | 1 | 0 | 80.0% (37.6-99.0) |
| Drugs | 0 | 0 | 0 | 0 | |
| Documents | 0 | 0 | 0 | 0 | |
| Radiology reports | 0 | 0 | 0 | 0 | |
| **Combination query** | | | | | |
| Problem or joint aspirations or ≥2 other criteria | 180 | 168 | 9 | 3 | 93.3% (88.7-96.1) |

*Supplementary table 6: Positive predictive value according to each query and a combination of queries, among 180 inpatients PPV.*

| Query (outpatients) | Total | Gout | No gout | Equivocal | Positive predictive value (CI 95%) |
|---|---|---|---|---|---|
| **Present irrespective of other criteria** | | | | | |
| ICD-10-GM Codes | 31 | 30 | 1 | 0 | 96.8% (83.8-99.8) |
| Problem list | 66 | 63 | 3 | 0 | 95.5% (87.5-98.4) |
| Joint aspiration | 10 | 9 | 1 | 0 | 90.0% (59.6-99.5) |
| Drugs | 76 | 76 | 0 | 0 | 100.0% (95.2-100.0) |
| Documents | 73 | 67 | 5 | 1 | 91.8% (83.2-96.2) |
| Radiology reports | 14 | 10 | 3 | 1 | 71.4% (45.4-88.3) |
| **Present alone** | | | | | |
| ICD-10-GM Codes | 0 | 0 | 0 | 0 | |
| Problem list | 4 | 3 | 1 | 0 | 75.0% (30.1-98.7) |
| Joint aspiration | 1 | 0 | 1 | 0 | 0.0% (0.0-94.9) |
| Drugs | 0 | 0 | 0 | 0 | |
| Documents | 0 | 0 | 0 | 0 | |
| Radiology reports | 0 | 0 | 0 | 0 | |
| **Combination query** | | | | | |
| Problem or joint aspirations or ≥2 other criteria | 82 | 74 | 7 | 1 | 90.2% (81.9-95.0) |

*Supplementary table 7: Positive predictive value according to each query and a combination of queries, among 82 outpatients PPV. ICD-10-GM codes are not available for inpatient.*

| Risk factor / Characteristics | Overall | Alcohol | MI | CKD | MS | Uricemia |
|---|---|---|---|---|---|---|
| Total number | 15646 | 3249 | 4818 | 4188 | 1906 | 1485 |
| Age (mean (SD)) | 73.09 (13.2) | 65.80 (13.2) | 73.68 (13.0) | 79.33 (11.1) | 71.75 (10.8) | 71.22 (13.6) |
| BMI (mean (SD)) | 33.78 (28.7) | 32.56 (25.9) | 32.85 (29.2) | 33.69 (29.7) | 38.80 (32.8) | 33.27 (22.6) |
| Number of death (%) at 31.12.2022 | 6567 (42.0) | 1051 (32.3) | 1882 (39.1) | 2314 (55.3) | 622 (32.6) | 698 (47.0) |
| Detected by the register (%) | 2588 (16.5) | 362 (11.1) | 524 (10.9) | 817 (19.5) | 287 (15.1) | 598 (40.3) |

*Supplementary Table 8: Absolute number of patients with risk factor for gout, their characteristics and number of patients detected by our algorithm detecting gout patients. MI: myocardial infarction. CKD: chronic kidney disease. MS: Metabolic syndrome. BMI: body mass index*

| Criterion present alone | Total | Gout | No gout | Equivocal | Negative predictive value (CI 95%) |
|---|---|---|---|---|---|
| Uricaemia > 500 µmol/l | 22 | 6 | 16 | 0 | 72.7% (51.8 to 86.8) |
| Chronic kidney disease | 86 | 5 | 79 | 2 | 91.9% (84.1 to 96.0) |
| Metabolic syndrome | 39 | 3 | 36 | 0 | 92.3% (79.7 to 97.3) |
| Myocardial infarction | 128 | 1 | 126 | 1 | 98.4% (94.5 to 99.6) |
| Deleterious use of alcohol | 107 | 2 | 104 | 1 | 97.2% (92.1 to 99.0) |

*Supplementary table 9 : Absolute numbers of patients and negative predictive value (NPV) of each criterion alone (a patient is detected only by the criterion). NPV was calculated by considering gout present versus gout absent or equivocal meaning equivocal cases were classified as gout in the NPV analysis.*

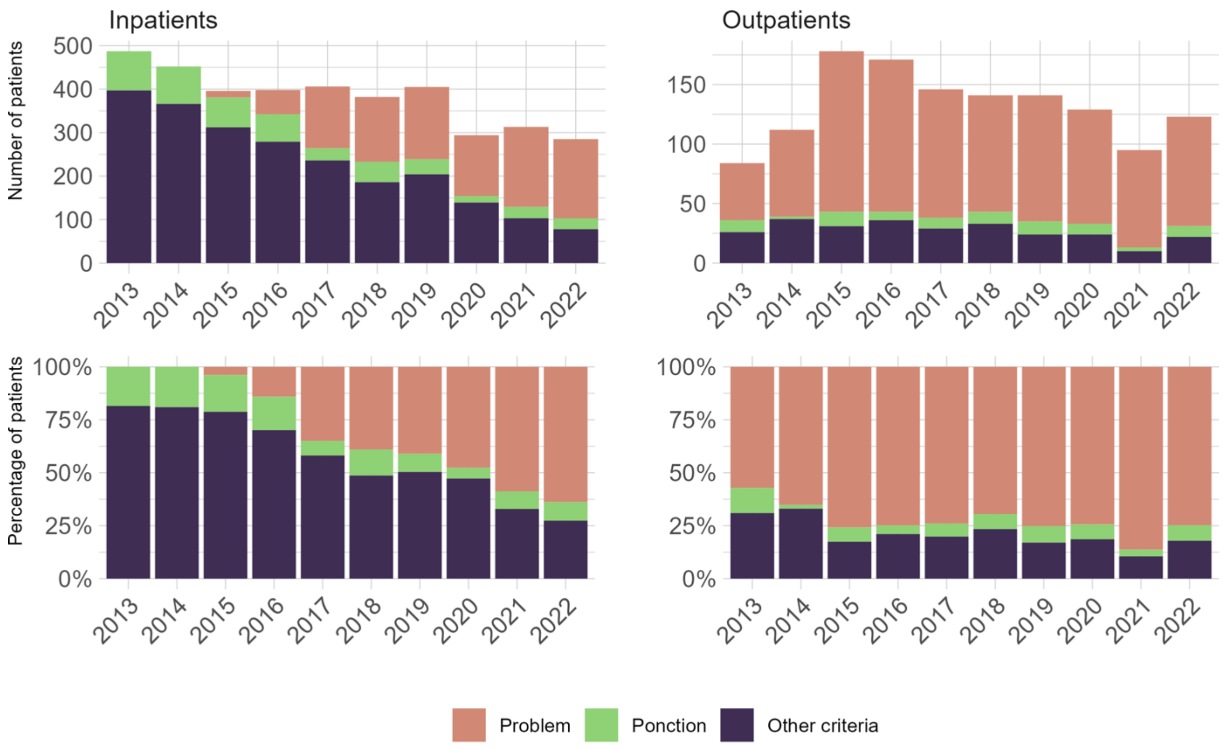


*Supplementary figure 2: Evolution of gout diagnostic according to the criterion or combination of criteria to establish the diagnosis at first detection in the register (or second in case of query combination). Other criteria correspond to a combination of ≥2 criterion among the documents, ICD-10-GM codes, imaging reports and drugs queries.*
